# Functional and Structural Neuroplasticity of Somatosensory System in Hemiplegic Cerebral Palsy

**DOI:** 10.1101/2025.08.14.25333613

**Authors:** Sadra Shahdadian, Yanlong Song, Alireza Vaysi, Setu Shiroya, Haden Ray, Margherita A. G. Matarrese, Madhan Bosemani, Fernando Acosta, Brooke Kimbrell, Warren Marks, Christos Papadelis

## Abstract

Children with hemiplegic cerebral palsy (HCP) exhibit diverse sensory deficits linked to varied atypical brain reorganization patterns. The underlying mechanisms of these patterns remain poorly understood. Using multimodal neuroimaging, we associate functional and structural neuroplasticity in the somatosensory system of children with HCP with lesion type and behavioral outcome. We hypothesize that different lesion types produce distinct patterns of neuroplasticity, which correlate with varying levels of sensory and motor behavioral deficits.

We examined 22 children with HCP (seven with cortical-subcortical (CSC), 12 with periventricular (PV), and three with other lesions), and 24 age-matched neurotypical controls. We assessed the somatosensory outcomes through touch sensitivity and two-point discrimination measures, and motor outcomes through movement range, accuracy, dexterity, and fluency measures. We quantified the integrity of ascending sensory (ASF) and somatosensory commissural (SCF) fibers with diffusion-weighted MRI and measured cortical responses to haptic stimulation with magnetoencephalography and high-density electroencephalography. Children with CSC lesions demonstrated higher sensory thresholds and poorer movement quality in the paretic hand compared to the PV lesion (*P <* 0.05) and control group (*P <* 0.01). Functional imaging revealed suppressed primary (S1) and secondary (S2) somatosensory cortical activations (*P <* 0.01) and prolonged S1 latency (*P <* 0.05) in the more affected hemisphere (MA) in response to paretic hand stimulation. These children also exhibited more pronounced disruption of interhemispheric functional balance (*P <* 0.05). Tractography showed greater microstructural damage—increased mean (*P <* 0.01), axial (*P <* 0.01), and radial (*P <* 0.01) diffusivities— in ASF and SCF in the MA of the CSC group. In contrast, the PV group showed unilateral damage to ASF (*P <* 0.01) and bilateral damage to SCF (*P <* 0.05). More severe diffusion abnormalities were associated with lower cortical amplitudes (*Rho*= –0.58 to – 0.78, *P <* 0.05), delayed latencies (*Rho*= 0.69, *P <* 0.05), and decreased lateralization (*Rho*= – 0.62 to –0.72, *P <* 0.05). Moderate to strong correlations were observed between cortical amplitudes and touch sensitivity (*Rho*= –0.56 to – 0.60, *P <* 0.05) and discriminability (*Rho*= – 0.77, *P <* 0.01).

Our findings show that children with HCP exhibit distinct structural and functional neuroplasticity patterns based on lesion type. CSC lesions lead to more extensive somatosensory damage, while PV lesions are linked to greater neuroplastic potential. These insights deepen our understanding of neuroplasticity mechanisms and highlight targets for optimizing therapeutic interventions.

## Introduction

Cerebral palsy encompasses a variety of neurodevelopmental disorders, mainly manifesting as movement and postural abnormalities, often associated with sensory deficits.^1–4^ The most predominant cerebral palsy form is hemiplegic cerebral palsy (HCP),^5,6^ which is characterized by heterogeneous levels of motor and somatosensory deficits on one side of the body, often affecting the hand functions and bimanual coordination.^7^ The severity of these deficits is influenced by several factors, including the underlying etiology, timing of the insult, and the extent to which the affected physiological system(s) respond to them. HCP is often caused by a non-progressive brain insult that occurs unilaterally during the perinatal period.^8^ The brain’s response to this insult is adaptive neuroplasticity through the formation of new neural connections.^9–14^ The extent and effectiveness of this neuroplasticity vary widely among individuals, since it is influenced by the timing and size of the lesion as well as the involved circuit(s).^15,16^ Despite the progress made to date, our understanding of adaptive neuroplasticity, particularly in the somatosensory system, remains limited, and the neural correlates of various neuroplasticity patterns remain unknown.

We currently lack a complete understanding of how etiology is associated with functional and structural neuroplasticity as well as behavioral outcomes. This lack of knowledge has led to generic, low-impact therapy approaches that fail to augment the individual’s neuroplasticity potential. Thus, there is an unmet need for individualized neuroplasticity mapping, especially for children with severe somatosensory deficits who commonly exhibit higher levels of hand impairments.^11^ Aiming to link the sensorimotor deficits with etiology, previous MRI studies categorized most children with HCP into either having periventricular (PV) or cortical-subcortical (CSC) lesions.^17,18^ PV lesions mainly occur in the early third trimester of gestation and are characterized by white matter volume loss and compensatory ventricular dilation.^19,20^ Contrarily, most CSC lesions typically occur in the late third trimester and postnatal period;^21^ these lesions cause volume loss of the cortical and subcortical gray matter, as well as the adjacent white matter.^22^ Children with CSC lesions experience more pronounced upper extremity impairments, particularly in the somatosensory function, than those with PV lesions.^11,18,23,24^ These differences in behavioral outcomes may be determined partially by the timing of the insult. For instance, the occurrence of PV insults before the development of ascending somatosensory fibers (ASF) usually leads to these fibers bypassing the lesion and reaching the spared cortex. In contrast, CSC lesions commonly damage the already developed ASF and somatosensory cortices that disrupt the transfer and processing of afferent information, respectively.^17,25^ To underscore the effect of perinatal brain insult on white matter fibers, previous diffusion-weighted imaging (DWI) studies have examined the microstructural integrity of ASF and found indications of damage within the more affected (MA) hemisphere.^11,13,14,18^ Other DWI studies have shown that microstructural damages are also extended to the association and commissural fibers.^14,26^ Yet, DWI studies do not offer insights on possible alterations in the functionality of the underlying somatosensory circuitry.

Functional neuroimaging methods, such as event-related potentials (ERP) and fields (ERF), provide critical insights into the sensorimotor circuit functionality. By employing these techniques, our group and others have shown alterations in the somatosensory evoked potentials/fields (SEP/SEF) of children with HCP, such as changes in amplitude, somatotopy, and phase synchronization between the primary (S1) and secondary (S2) somatosensory cortices.^12,13,27,28^ These findings reveal maladaptive changes in the function of different nodes in the somatosensory circuit (e.g., S1 and S2),^12,13,28^ and distorted information transmission between them.^28^ Stratifying different lesion types and their distinct effect on this circuit’s functionality may explain variability in behavioral outcomes and aid in tailoring future rehabilitation approaches.^25,29^ By employing multimodal neuroimaging, we aim to identify atypical patterns of functional and structural somatosensory reorganization in the brain of children with HCP and their association with the lesion type and behavioral outcome. We hypothesize that different lesion types produce distinct neuroplasticity patterns, which correlate with varying levels of somatosensory and motor behavioral deficits. To assess the functional alterations in the somatosensory circuit and interhemispheric balance, we recorded simultaneously ERPs and ERFs in response to haptic stimulation and localized S1 and S2 through electromagnetic source imaging (EMSI)^30^ in a cohort of children with HCP and a group of neurotypically developing controls. To assess the microstructural integrity of ASF and somatosensory commissural fibers (SCF), we used functionally guided DWI tractography. We classified most HCP children in our cohort into two groups (i.e., CSC and PV groups) based on lesion type. We then examined associations between functional neuroplasticity, structural alterations in the ASF and SCF, and sensorimotor behavioral outcomes across groups.

## Materials and methods

### Participants

We recruited 22 children with HCP (15 male, 10.8 ± 3.2 years) and 24 typically developing (TD) children (13 male, 11.0 ± 3.4 years). Children with HCP were recruited from the movement disorders clinic at Cook Children’s Medical Center (CCMC), Fort Worth, TX. TD children were recruited from the local community. The inclusion criteria for the HCP participants were the following: (1) evaluation by a pediatric neurologist, neonatal developmental specialist, or neonatologist with an HCP diagnosis; (2) classified as high-functioning (GMFCS I or II); and (3) age-appropriate understanding of study procedures. The exclusion criteria were as follows: (1) psychoactive or myorelaxant medication during study procedures; (2) genetic syndrome diagnosis; (3) history of trauma or brain surgery; (4) inability to sit still; and (5) metal implants or baclofen pumps in the body. TD children had no neurological disorder or brain injury. The Institutional Review Board at Cook Children’s Health Care System approved this study (2019-068; PI: C. Papadelis).

### Behavioral assessment

#### Touch sensitivity and discrimination

We tested touch sensitivity on the thumb (D1), middle finger (D3), and pinky (D5) of both hands using Semmes-Weinstein monofilaments^31^ (Figure 1A). The monofilaments were applied in a random order while the participant had his/her eyes closed. For each stimulus, participants reported perception. Each monofilament was applied three times per finger. A touch sensitivity threshold was determined for each finger as the lowest weight perceived twice.

**Figure 1.**
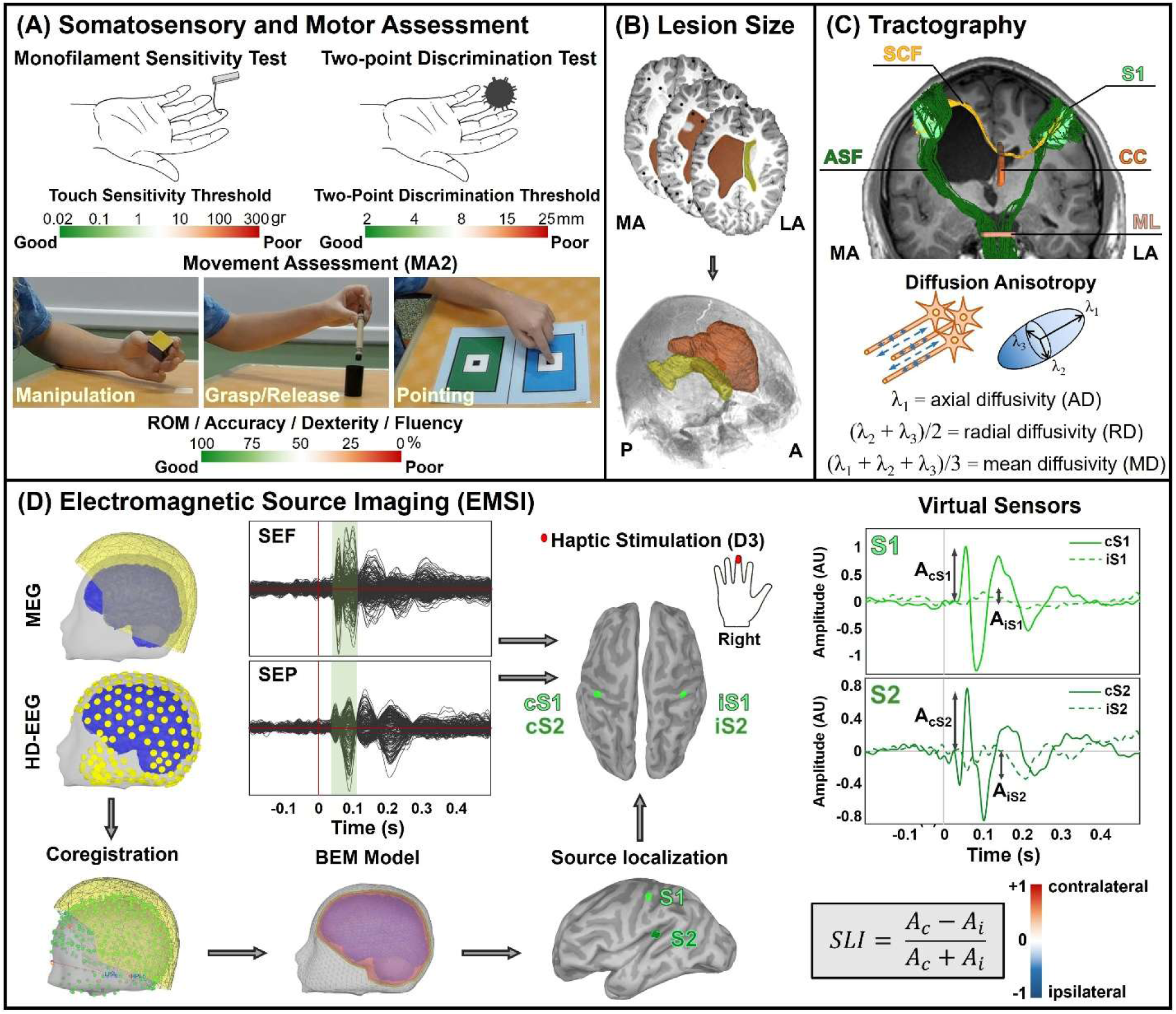
Overview of data collection and analysis. **(A)** Somatosensory and motor assessments, including monofilament sensitivity, two-point discrimination, and movement quality evaluation. **(B)** Reconstruction of bilateral ventricular volumes from T1-weighted MRI of a participant with a periventricular (PV) lesion (#9). Lesion/ventricular dilation size quantified by subtracting ventricular volume in the less affected (LA) from the more affected (MA) hemisphere. **(C)** Tractography derived from diffusion-weighted imaging (DWI). Anatomically defined (corpus callosum [CC], medial lemniscus [ML]) and functionally guided (primary somatosensory cortex [S1]) regions of interest were used to isolate ascending sensory (ASF) and somatosensory commissural fibers (SCF). Diffusion metrics—axial diffusivity (AD), radial diffusivity (RD), and mean diffusivity (MD)—were extracted from each tract. **(D)** Electromagnetic source imaging pipeline. Magnetoencephalography (MEG; 306 sensors) and high-density EEG (HD-EEG; 256 channels) data extracted somatosensory evoked fields (SEF) and potentials (SEP) at the sensor level. MEG and EEG sensor locations coregistered onto the subject’s 3D head and cortical surfaces. A realistic boundary element method (BEM) head model constructed from the subject’s T1-weighted MRI. The inverse model localized activity in bilateral primary (S1) and secondary (S2) somatosensory cortices from haptic stimulation of the middle finger (D3). Virtual sensors in bilateral S1 and S2 provided cortical activity time series. Amplitude and latency of peak activity computed in both S1 and S2. Somatosensory lateralization index (SLI) calculated for S1 and S2 to assess interhemispheric balance between contralateral and ipsilateral responses. A = amplitude; c = contralateral; i = ipsilateral; MA2 = Melbourne Assessment 2.

We also tested static two-point discrimination using the Touch Test® Two-Point discriminator^32^ (Exacta™ Evaluation Kit; North Coast Medical Inc., CA). The discrimination thresholds were determined on D1, D3, and D5 of both hands three times by randomly applying either one- or two-point stimulus to fingertips. The participant was asked to state whether he/she perceived one- or two-point stimulus. A discrimination threshold was determined for each finger as the lowest distance perceived twice.

#### Movement quality

We evaluated unilateral upper-extremity function, including range of motion (ROM), accuracy, dexterity, and fluency, using the Melbourne Assessment 2 (MA2)^33^ (Figure 1A). Children were given various items to manipulate and were scored differently related to each movement element. A total of 30 item scores was used. Scoring was conducted by two double-blinded researchers (S.S. and E.A.) and was completed across the 30 score items using a 3, 4, or 5 scale and individually defined scoring criteria. Item scores relating to each element measured were summed within the corresponding sub-scale.

#### MRI/DWI acquisition

The imaging protocol included structural and DWI sequences collected at Radiology of CCMC. We obtained T1-weighted structural MRI scans using a Siemens Skyra 3T MR scanner and a 10-channel head coil with 3D magnetization-prepared rapid-acquisition gradient-echo sequence with parameters: echo time = 3.22 ms, inversion time = 450 ms, repetition time = 8.21 ms, flip angle = 15 degrees, field of view = 24 cm, matrix size = 256 × 256, 184 slices, and resolution = 1.0 × 1.0 × 1.0 mm³. DWI employed a multi-shell diffusion scheme (b-values: 2000 s/mm²) with diffusion sampling directions of 30 and 60, respectively. The in-plane resolution was 2.04918 mm, and the slice thickness was 2 mm.

#### Lesion type, affected hemispheres, and structures

A pediatric neuroradiologist (M.B.) reviewed the MRI scans. Together with a pediatric neurologist (W.M.), they identified the underlying HCP lesion type, as well as the affected hemisphere and cortical/sub-cortical structures. For all HCP participants, the hemisphere with the most pronounced structural abnormalities was labeled as MA. PH was contralateral to the MA hemisphere in all HCP participants. Clinical characteristics of HCP participants are in Table 1. In TD participants, we marked the contralateral hemisphere to the dominant hand (DH) as the dominant hemisphere (D).

**Table 1.** demographics and clinical characteristics of hemiplegic cerebral palsy participants.

| # | Gender | Affected body side | MRI findings (side) | Brain structures affected | Lesion type | GMFCS | MACS | Functional neuroimaging |
| --- | --- | --- | --- | --- | --- | --- | --- | --- |
| 1 | M | R | MCA stroke (L) | Cortex, WM | CSC | 2 | 2 | MEG, HD-EEG |
| 2 | M | R | MCA stroke (L) | Cortex, WM, BG, thalamus | CSC | 1 | 5 | MEG |
| 3 | F | L | IVH (R: IV, L: II) | Cortex, WM, CC, thalamus | PV | 1 | 2 | MEG, HD-EEG |
| 4 | M | R | HIE (L) | WM, CC, BG, thalamus | PV | 2 | 1 | MEG, HD-EEG |
| 5 | M | R | MCA stroke (L) | Cortex, WM, thalamus | CSC | 2 | 1 | MEG, HD-EEG |
| 6 | M | L | IVH (R: IV) | WM, CC, BG, thalamus | PV | 2 | 2 | MEG, HD-EEG |
| 7 | M | L | MCA stroke (R) | Cortex, WM, BG, thalamus | CSC | 2 | 5 | MEG, HD-EEG |
| 8 | F | R | PVL (L>R) | WM, BG, thalamus | PV | 1 | 1 | MEG, HD-EEG |
| 9 | M | R | IVH (L: IV) | WM, BG, CC | PV | 2 | 3 | MEG, HD-EEG |
| 10 | F | L | Polymicrogyria (R) | Cortex | other | 1 | 3 | MEG |
| 11 | M | L | Polymicrogyria pachygyria (R) | Cortex | other | 2 | 2 | MEG, HD-EEG |
| 12 | M | R | MCA stroke (L) | Cortex, WM, thalamus | CSC | 1 | 3 | MEG, HD-EEG |
| 13 | F | R | MCA stroke (L) | Cortex, WM, CC, BG, thalamus | CSC | 2 | 3 | MEG |
| 14 | M | R | MCA stroke (L) | Cortex, WM, CC, BG, thalamus | CSC | 2 | 5 | MEG, HD-EEG |
| 15 | M | R | IVH (L: II) | WM | PV | 1 | 1 | MEG, HD-EEG |
| 16 | F | R | Schizencephaly, polymicrogyria | Cortex, WM | other | 2 | 3 | MEG, HD-EEG |
| 17 | M | L | IVH (L: IV) | WM, CC, BG, thalamus | PV | 1 | 3 | MEG, HD-EEG |
| 18 | M | R | IVH (L: II) | WM, CC | PV | 2 | 2 | MEG, HD-EEG |
| 19 | M | L | HIE - PVL (L>R) | WM | PV | 1 | 2 | MEG |
| 20 | F | L | PVL (R>L) | WM | PV | 2 | 1 | MEG, HD-EEG |
| 21 | F | R | Centrum semiovale gliosis (L) | WM | PV | 2 | 1 | MEG |
| 22 | M | L | Periventricular WM gliosis(R) | WM | PV | 1 | 1 | MEG, HD-EEG |

M = male; F = female; L = left; R = right; MCA = middle cerebral artery; IVH = intraventricular hemorrhage; HIE = hypoxic-ischemic encephalopathy; PVL = periventricular leukomalacia; WM = white matter; CC = corpus callosum; BG = basal ganglia; CSC = cortical-subcortical; PV = periventricular; GMFCS = gross motor function classification system; MACS = manual ability classification system; MEG = magnetoencephalography; EEG = electroencephalography.

#### Calculation of lesion/ventricular dilation size

We determined the lesion/ventricular dilation size from the axial T1-weighted MRI and verified it in coronal/sagittal views. The lesion borders were manually marked (A.V.) with MATLAB® Volume Segmenter Toolbox. The corresponding lesion/dilated ventricle volume was then computed (Figure 1B). Lesion volume included regions of gray matter loss and ex-vacuo ventricular dilation secondary to white matter volume loss. To account for baseline ventricle size, we subtracted the less affected (LA) hemisphere’s ventricular volume from the total volume of the lesion and dilated ventricle in the MA hemisphere.

#### Acquisition of ERP/ERF

We performed simultaneous magnetoencephalography (MEG) and high-density electroencephalography (HD-EEG) recordings inside a single-layer magnetically shielded room (MSR) located at suite Radiology of CCMC. MEG recordings were performed with a whole head 306 sensor system (VectorView, Elekta Neuromag, Helsinki, Finland). HD-EEG recordings were performed with the Geodesic EEG System 400 (Magstim EGI Inc., USA) that uses nets with either 128 or 256 channels (depending on participant’s head size). Before data acquisition, we placed five head position indicator coils on child’s head, and used a digitizer (Fastrak Polhemus, USA) to map head cardinal landmarks and ∼500 scalp points into digital 3D coordinates. This enabled alignment of MEG sensors with participant’s MRI for source localization. We also recorded ECG and EOG signals for artifact removal during preprocessing. Before MSR entry, we positioned the EEG net on the participant’s head and checked the electrode impedances to ensure adequate conductivity (impedance < 50 KΩ). To determine the electrode positions with respect to the participant’s head, we used the GeoScan Sensor Digitization system (Magstim EGI Inc., USA). When simultaneous MEG and HD-EEG were not feasible, we collected only MEG data (Table 1). For SEF and SEP recordings, we delivered compressed air bursts to middle fingertips (D3) through a pneumatic stimulation system (AIRSTIM, San Diego Instruments, USA). We applied stimulations asynchronously with a 1.5 ± 0.5-s inter-stimulus interval following a pseudorandom sequence (400 stimuli per finger). Participants kept their arms motionless on a tray and watched cartoons to minimize movement artifacts, while preserving SEF and SEP characteristics.^34^

#### Electromagnetic data analysis

We processed the raw MEG signals using MaxFilter (MEGIN, Finland) to reduce noise and correct for head motions. Furthermore, an in-house algorithm^30^ spatially aligned and temporally synchronized the MEG/HD-EEG data. We then performed band-pass (1-100 Hz) and notch (60 Hz) filters on MEG and HD-EEG data using *Brainstorm*.^35^ artifacts generated by heartbeats, eye movements and blinks, and muscle contractions were removed through SSP and ICA methods^36,37^ for MEG and HD-EEG data, respectively.

#### Source localization

To obtain SEF and SEP evoked by finger stimulation, we segmented continuous sensor data into event-locked trials of 700 ms (200 pre- and 500 ms post-stimulus). We calculated grand averages by averaging trials for each participant. Dynamic Statistical Parametric Mapping (dSPM)^38^ was employed to identify neural sources.^39^ We generated canonical surfaces (i.e., cortex, inner skull, outer skull, and scalp) from individual T1-weighted MRI data. We then co-registered MEG and HD-EEG signals by projecting digitized cranial locations onto scalp’s surface and estimated the forward models using the boundary element modeling (BEM) with *OpenMEEG*.^40^ We derived noise covariance matrices from pre-stimulus activity. Using *Brainstorm’s* default parameters, we computed the dSPM source activity for ∼15,000 cortical vertices, incorporating gradiometers, magnetometers, and HD-EEG channels. We identified peak cortical activities corresponding to S1 and S2, located in Brodmann’s areas 3/1/2 and 43, respectively. We extended the S1 cortical vertices to surrounding vertices for defining an area of ∼3 cm² and exported the coordinates as region of interest (ROI) for tractography (Figure 1D). Due to severe cortical damage in two participants from the CSC group, the locations of the first and second SER peaks in response to haptic stimulation of the PH did not align with the anatomical atlas. Therefore, we selected the vertices with peak amplitude on the MA side as S1 and S2.

#### Somatosensory lateralization index

To assess functional interhemispheric balance between lateral S1 and S2, we computed the cortical somatosensory lateralization index (*SLI*) for the TD, CSC, and PV groups in response to each hand. We placed virtual sensors on bilateral EMSI-guided S1 and S2 cortical vertices to extract somatosensory evoked responses (SER) during lateral finger stimulation (Figure 1D). We baseline-normalized the maximum activation value in each area and its contralateral counterpart and calculated SLI as:^41^

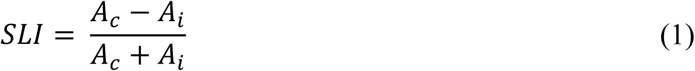

where *A_c_* and *A_i_* represent amplitude of SER in contralateral and ipsilateral hemispheres, respectively. *SLI* ranges from -1 (ipsilateral activity) to +1 (contralateral activity).

#### ROI-based tractography

We performed DWI preprocessing and tractography using *DSI-Studio*. Our preprocessing steps included visual quality inspection, motion correction, and the creation of a white matter mask. We performed fiber tracking using Generalized Q-sampling Imaging (GQI),^42^ which allows for the identification of complex white matter configurations, including crossing fibers. We set the threshold for tract angle to 60 degrees, step sizes from 0.5 to 1.5 voxels, and track lengths from 30 to 200 mm. We isolated a total of 3000 tracts and applied topology-informed pruning^43^ over 16 iterations to eliminate false connections.

We loaded primary somatosensory areas derived from EMSI into *DSI-Studio* and identified the ASF by setting S1 and the medial lemniscus as ROIs (Figure 1C). We also defined SCF, which pass through the corpus callosum (CC), by setting unilateral S1 and the CC as ROIs. We then trimmed the identified SCFs to retain only the segments located in the hemisphere ipsilateral to the selected S1. In addition, GQI allowed us to estimate diffusivity parameters [i.e., mean diffusivity (MD), axial diffusivity (AD), and radial diffusivity (RD)] of the reconstructed fibers to assess white matter microstructural integrity.

#### Statistical analysis

An a priori power analysis using G*Power software determined that a total of 42 participants is needed to detect lesion-specific effects with medium effect size (Cohen’s f = 0.5), alpha = 0.05, and power = 0.8. To identify lesion-specific patterns of behavioral, neurophysiological, and microstructural alterations in the somatosensory network, we performed statistical analyses on data from TD participants and children with PV and CSC lesions. We used Wilcoxon signed rank tests to compare behavioral (i.e., sensitivity, discriminability, and MA2 parameters), functional (i.e., S1 and S2 amplitude, latency, and SLI), and microstructural (i.e., MD, AD, and RD) measures within hands/hemispheres of each group (i.e., TD, CSC, and PV groups). We also employed Kruskal-Wallis tests to compare these measures among two sets of data: (i) non-dominant hand (NDH)/hemisphere (ND) of TD vs paretic hand (PH)/MA of CSC and PV group; (ii) DH/D of TD vs non-paretic hand (NPH)/LA of CSC and PV groups. The significance level was set at 0.05. For post hoc testing, we performed pairwise Mann-Whitney U tests with false discovery rate (FDR) correction for multiple comparisons.

We used Spearman correlations to identify significant associations among behavioral outcomes, functional measures, and structural (i.e., lesion size, MD, AD, and RD) measures. In these analyses, we aimed to examine associations between the: (i) functional and behavioral measures; (ii) structural and behavioral measures; and (iii) structural and functional measures. To account for uneven sample sizes, we performed two sets of correlation analyses: one that included all conditions (i.e., both hands/hemispheres of the TD, CSC, and PV groups), and another that included only the PH or the MA hemisphere of the CSC and PV groups. The significance level was set at 0.05. To correct for multiple comparisons, we performed FDR correction. All statistical analyses were conducted using R® version 4.5.0.

## Results

### Behavioral assessment

#### Touch sensitivity

Finger-level analysis revealed elevated touch sensitivity thresholds in the MA hand of the CSC group compared to NDH of TD participants (D1, D5: *P <* 0.01; D3: *P <* 0.05) and the PH of the PV group (*P <* 0.05) (Figure 2A). With pooled data from all three fingers, thresholds remained significantly higher in the MA hand of the CSC group compared to its LA (*P <* 0.001), PH of the PV group (*P <* 0.001), and NDH of TD group (*P <* 0.001). The PH of the PV group also showed higher thresholds than its non-paretic counterpart and NDH of TD group (*P <* 0.05).

**Figure 2.**
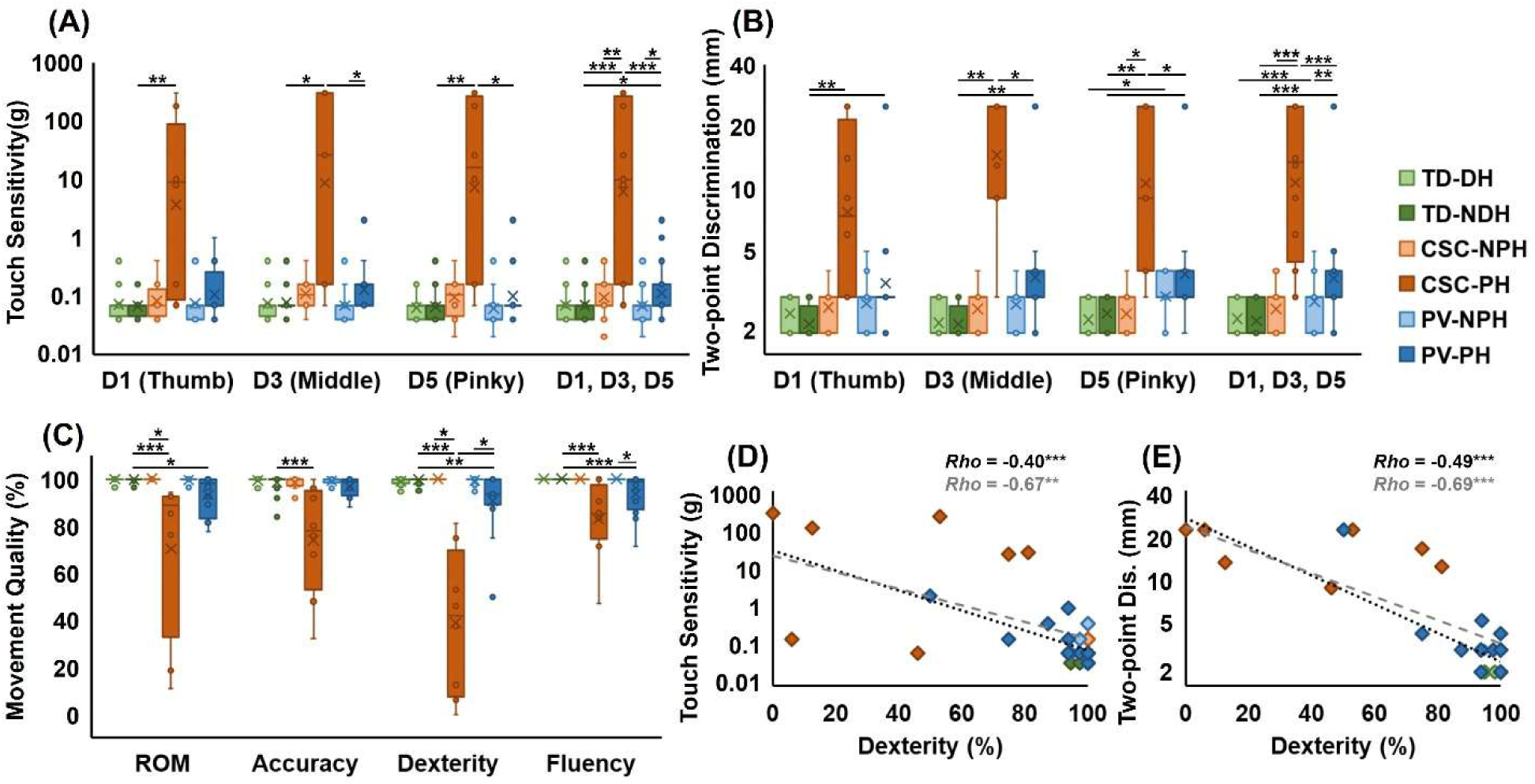
Behavioral assessment results. **(A)** Box-and-whisker plots of touch sensitivity thresholds (g, log scale) at thumb (D1), middle finger (D3), pinky (D5) and the pooled D1–D5 in the dominant (TD-DH) and non-dominant (TD-NDH) hands of typically developing (TD) controls and the non-paretic (NPH) and paretic (PH) hands of children with periventricular (PV) or cortical–subcortical (CSC) lesions. **(B)** Static two-point discrimination thresholds (mm) in the same groups and digits. **(C)** Melbourne Assessment 2 sub-scores (%) for range of motion (ROM), accuracy, dexterity and fluency in the same groups. **(D, E)** Spearman correlations between dexterity sub-score and touch sensitivity **(D)** or two-point discrimination **(E)** in the PH, shown across all participants (black) and across PH of HCP participants only (gray). Boxes indicate median and interquartile range, whiskers range, × denotes mean. *p<0.05, **p<0.01, ***p<0.001 derived from Wilcoxon signed rank tests for within subject, and Kruskal-Wallis and Mann-Whitney U tests for between subject comparisons (FDR corrected for multiple comparisons).

#### Two-point discrimination

Two-point discrimination thresholds were elevated in the PH of the CSC group compared to NDH of TD group (*P <* 0.01), as well as its NPH (D5: *P <* 0.05; Figure 2B). Within the PV group, significant differences were observed between both hands and TD group (PH: D1, D3: *P <* 0.01; D5: *P <* 0.05; NPH: D5: *P <* 0.05). Pooled three fingers, data demonstrated elevated thresholds in the PHs of CSC group and both hands of PV group (*P <* 0.001), relative to TD children. However, the PH of the PV group still showed lower thresholds than the CSC group (*P <* 0.001).

#### Movement assessment

The PH of the CSC group exhibited reduced performance in all four domains of ROM, accuracy, dexterity, and fluency, compared to its NPH and NDH of TD children (*P <* 0.001; Figure 2C). The PH of the PV group also showed decreased ROM (*P <* 0.05), dexterity (*P <* 0.01), and fluency (*P <* 0.001), relative to NDH of TD group. Strong negative correlations were observed between dexterity and hand-averaged touch sensitivity (*Rho* = –0.67, *P <* 0.01), as well as two-point discrimination thresholds (*Rho* = –0.69, *P <* 0.001) in the PHs of children with CSC and PV lesions (Figure 2D and 2E).

#### Anatomical alterations

All children with HCP had predominantly unilateral structural defects. Among the 22 children with HCP, 7 had CSC lesions (six male, 11.0 ± 3.2 years), 12 had PV lesions (eight male, 10.3 ± 3.4 years), and three had other lesions (one male, 12 ± 1.4 years). MRI review indicated white matter damage in all children with PV and CSC lesions. Cortical gray matter injury was seen in all CSC cases (7 of 7), but only in one PV case (1 of 12). CC thinning, primarily in the posterior body, was more frequently observed in the PV group (6 of 12) than in the CSC group (2 of 7). Basal ganglia and thalamic injuries were more common in CSC participants (basal ganglia: 4 of 7; thalamus: 6 of 7) compared to the PV group (basal ganglia: 5 of 12; thalamus: 5 of 12). MRI of TD children showed no structural abnormalities.

Figure 3A shows examples of areas identified on MRIs of CSC and PV lesions. T1-weighted MRI analysis revealed larger lesion volumes in the CSC group compared to the PV group (*P <* 0.05; Figure 3B). Lesion/ventricular dilation size showed a positive correlation with PH touch sensitivity (*Rho* = 0.64, *P <* 0.01; Figure 3C) and two-point discrimination thresholds (*Rho* = 0.52, *P <* 0.05), indicating that larger lesions are associated with more severe sensory impairments (Figure 3D).

**Figure 3.**
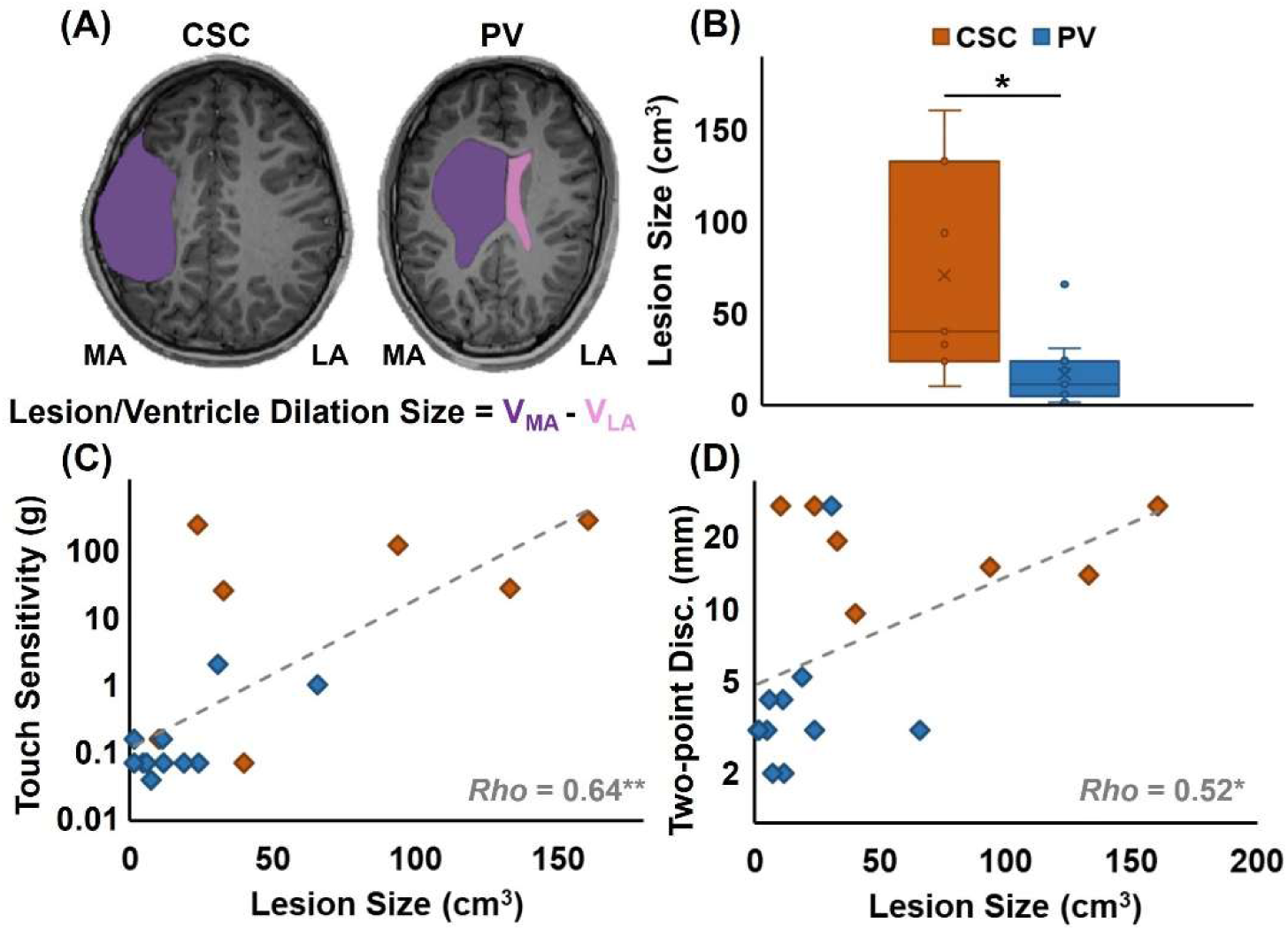
Lesion/ventricular dilation volumes and their relation to sensory thresholds. **(A)** Representative axial T1-weighted MRIs from CSC and PV groups with manual segmentations of enlarged ventricles/lesion cavities in the more affected (MA; purple) and less affected (LA; pink) hemispheres. **(B)** Box-and-whisker plot of lesion/ventricular dilation volumes (cm^3^) in CSC and PV groups. **(C, D)** Spearman correlations between lesion/ventricular dilation size in the MA hemisphere and touch sensitivity threshold (g; **C**) or two-point discrimination (mm; **D**) in the PH. *p<0.05, **p<0.01 derived from Mann-Whitney U test and Spearman Correlation.

#### SER features

Stimulation of the PH in the CSC group resulted in suppressed contralateral SER amplitudes in S1 and S2 (*P <* 0.01) compared to NDH of TD controls and PH of PV group (Figure 4A). Additionally, the NPHs of both CSC and PV groups showed decreased contralesional S1 activation relative to the TD group (*P <* 0.05). The CSC group displayed prolonged contralateral S1 latency in response to stimulation of the PH compared to their NPH (*P <* 0.05; Figure 4B). Stimulation of the PH in the CSC group resulted in reduced S1 lateralization (*P <* 0.05) compared to TD participants (Figure 4C). Additionally, S2 activation was higher in the NPH compared to the PH within the CSC group (*P <* 0.05).

**Figure 4.**
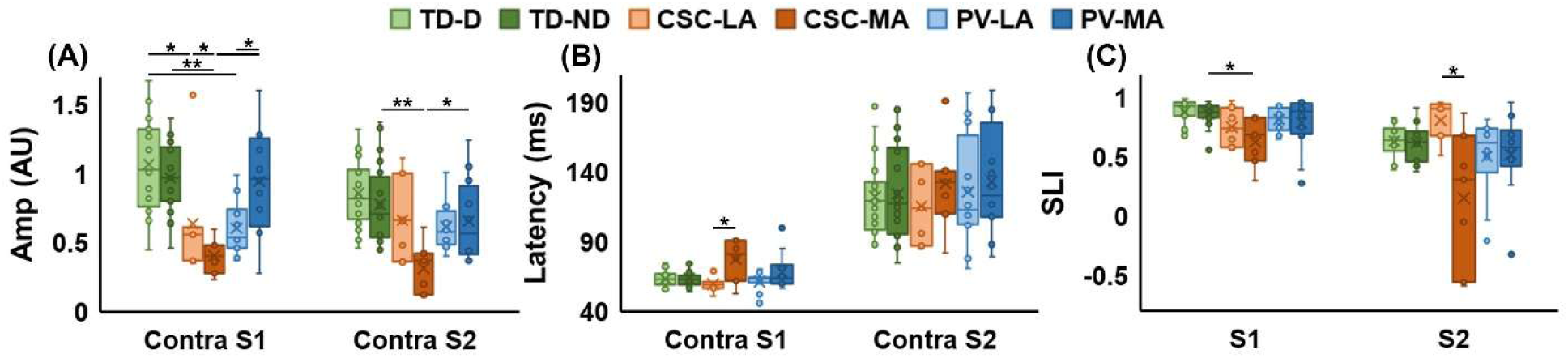
Somatosensory evoked potential amplitudes, latencies and lateralization. Box-and-whisker plots of contralateral S1 and S2 peak amplitude (arbitrary units; **A**), peak latency (ms; **B**) and somatosensory lateralization index (SLI; **C**) in response to haptic stimulation of middle finger of dominant (TD-D) and non-dominant (TD-ND) hemispheres of TD controls and the less affected (LA) and more affected (MA) hemispheres of CSC and PV groups. *p<0.05, **p<0.01, ***p<0.001 derived from Wilcoxon signed rank tests for within subject, and Kruskal-Wallis and Mann-Whitney U tests for between subject comparisons (FDR corrected for multiple comparisons).

### Tractography

#### Ascending sensory fibers (ASF)

MD was elevated in the MA hemisphere of both CSC and PV groups when compared to their corresponding LA hemispheres (CSC: *P <* 0.05; PV: *P <* 0.01) and ND of the TD group (*P <* 0.01; Figure 5A). AD and RD showed similar differences among three groups (see Figure 5A). This observation reflects a completely unilateral microstructural damage to ASF in both lesion types with greater severity in CSC than PV cases (MD, RD: *P <* 0.01; AD: *P <* 0.05).

**Figure 5.**
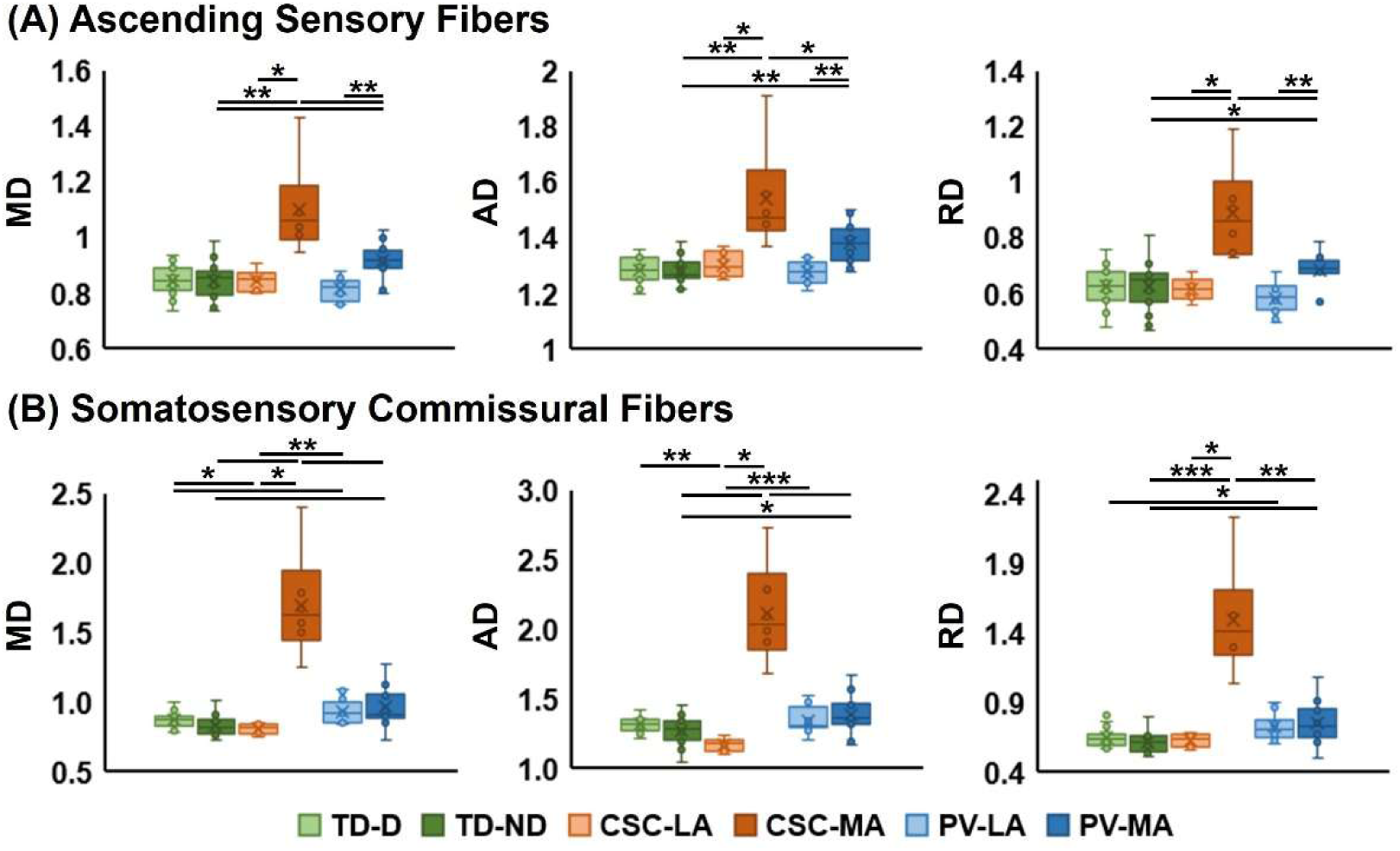
Diffusivity metrics in somatosensory white-matter tracts. Box-and-whisker plots of mean diffusivity (MD), axial diffusivity (AD) and radial diffusivity (RD) in ascending sensory fibers (ASF; **A**) and somatosensory commissural fibers (SCF; **B**) for the dominant (TD-D) and non-dominant (TD-ND) hemispheres of TD controls and the less affected (LA) and more affected (MA) hemispheres of CSC and PV groups. *p<0.05, **p<0.01, ***p<0.001 derived from Wilcoxon signed rank tests for within subject, and Kruskal-Wallis and Mann-Whitney U tests for between subject comparisons (FDR corrected for multiple comparisons).

#### Somatosensory commissural fiber (SCF)

MD was elevated in the MA hemisphere of CSC children compared to their LA hemisphere (*P <* 0.05) and ND hemisphere of TD group (*P <* 0.01; Figure 5B). The PV group showed increased MD in both hemispheres relative to the TD group (*P <* 0.05) and in the LA hemisphere compared to the LA hemisphere of the CSC group (*P <* 0.01). AD was also higher in the MA hemisphere of CSC participants when compared to its LA hemisphere (*P <* 0.05) and ND of TD children (*P <* 0.001). Interestingly, the AD in the CSC group’s LA hemisphere was lower than D hemispheres of the TD group (*P <* 0.01) and the LA of the PV group (*P <* 0.001). RD was higher in the CSC group’s MA hemisphere compared to its LA side (*P <* 0.05) and ND hemisphere of TD group (*P <* 0.001). The PV group also showed elevated RD bilaterally compared to the TD group (*P <* 0.05). These findings confirm SCF microstructural damage in the MA hemisphere of the CSC group and both hemispheres of the PV group, with greater severity in the CSC group (MD, RD: *P <* 0.01; AD: *P <* 0.001).

### Inter-modality associations

#### Functional measures - behavioral outcomes

Reduced S1 and S2 amplitudes were strongly associated with poorer tactile function. S1 amplitude correlated with touch sensitivity (*Rho* = –0.60, *p* < 0.05; Table 2) and two-point discrimination (*Rho* = – 0.77, *p* < 0.05). Prolonged latency in S1 correlated with higher touch thresholds (*Rho* = 0.63, *p* < 0.05). Lateralization indices in both S1 and S2 were negatively correlated with discrimination thresholds (S1 LI: *Rho* = –0.73, *p* < 0.05; S2 LI: *Rho* = –0.55, *p* < 0.05).

**Table 2.** Results for inter-modality structural, functional, and behavioral metric correlation.

| Metric #1 | Metric #2 | All subjects & sides |  | MA/PH of HCP Groups |  |
| --- | --- | --- | --- | --- | --- |
|  |  | Spearman's Rho | Significance | Spearman's Rho | Significance |
| Lesion size | Touch sensitivity | N/A | - | 0.64 | ** |
| Lesion size | 2-point discrimination | N/A | - | 0.52 | * |
| Contra S1 amp | Touch sensitivity | -0.31 | ** | -0.60 | ** |
| Contra S1 amp | 2-point discrimination | -0.50 | *** | -0.77 | ** |
| Contra S2 amp | Touch sensitivity | -0.24 | * | -0.49 | ** |
| Contra S2 amp | 2-point discrimination | -0.29 | ** | -0.43 | - |
| Contra S1 latency | Touch sensitivity | 0.17 | - | 0.63 | ** |
| S1 LI | Touch sensitivity | -0.18 | - | -0.47 | * |
| S1 LI | 2-point discrimination | -0.39 | *** | -0.73 | *** |
| S2 LI | Touch sensitivity | -0.09 | - | -0.54 | * |
| S2 LI | 2-point discrimination | -0.22 | * | -0.55 | * |
| ASF MD | Touch sensitivity | 0.26 | * | 0.72 | ** |
| ASF AD | Touch sensitivity | 0.34 | ** | 0.75 | *** |
| ASF RD | Touch sensitivity | 0.22 | - | 0.63 | ** |
| ASF MD | Contra S1 amp | -0.06 | - | -0.55 | * |
| ASF RD | Contra S1 amp | -0.02 | - | -0.50 | * |
| ASF MD | Contra S1 latency | 0.15 | - | 0.56 | * |
| ASF AD | Contra S1 latency | 0.21 | - | 0.69 | ** |
| ASF RD | Contra S1 latency | 0.13 | - | 0.53 | * |
| ASF MD | Contra S2 amp | -0.26 | * | -0.60 | * |
| ASF AD | Contra S2 amp | -0.40 | *** | -0.55 | * |
| ASF RD | Contra S2 amp | -0.17 | - | -0.54 | * |
| ASF MD | S2 LI | -0.20 | - | -0.71 | ** |
| ASF AD | S2 LI | -0.34 | ** | -0.64 | ** |
| ASF RD | S2 LI | -0.15 | - | -0.68 | ** |
| SCF MD | Touch sensitivity | 0.26 | * | 0.72 | ** |
| SCF AD | Touch sensitivity | 0.22 | * | 0.69 | ** |
| SCF RD | Touch sensitivity | 0.29 | * | 0.74 | *** |
| SCF MD | 2-point discrimination | 0.36 | ** | 0.63 | ** |
| SCF AD | 2-point discrimination | 0.39 | *** | 0.70 | ** |
| SCF RD | 2-point discrimination | 0.33 | ** | 0.65 | ** |
| SCF MD | Contra S1 amp | -0.18 | - | -0.77 | *** |
| SCF AD | Contra S1 amp | -0.17 | - | -0.78 | *** |
| SCF RD | Contra S1 amp | -0.22 | - | -0.76 | *** |
| SCF RD | Contra S1 latency | 0.15 | - | 0.52 | * |
| SCF MD | S1 LI | -0.18 | * | -0.67 | ** |
| SCF AD | S1 LI | -0.13 | - | -0.72 | ** |
| SCF RD | S1 LI | -0.25 | - | -0.66 | ** |
| SCF MD | Contra S2 amp | -0.22 | * | -0.66 | ** |
| SCF AD | Contra S2 amp | -0.14 | - | -0.58 | * |
| SCF RD | Contra S2 amp | -0.27 | * | -0.67 | ** |
| SCF MD | S2 LI | -0.37 | ** | -0.61 | ** |
| SCF AD | S2 LI | -0.34 | ** | -0.48 | * |
| SCF RD | S2 LI | -0.35 | ** | -0.62 | ** |

AD = axial diffusivity; amp = amplitude; ASF = ascending sensory fiber; HCP = hemiplegic cerebral palsy; LI = lateralization index; MA = more affected; MD = mean diffusivity; PH = paretic hand; RD = radial diffusivity; S1 = primary somatosensory cortex; S2 = secondary somatosensory cortex; SCF = somatosensory commissural fiber.
Statistically significant p-values obtained from Spearman's Rho are displayed; \*p < 0.05, \*\*p < 0.01, \*\*\*p < 0.001.

#### Microstructural measures - behavioral outcomes

For ASF, touch sensitivity was positively correlated with MD (*Rho* = 0.72, *p* < 0.05), AD (*Rho* = 0.75, *p* < 0.05), and RD (*Rho* = 0.63, *p* < 0.05). For SCF, MD, AD, and RD were strongly correlated with both behavioral measures—for example, RD with touch sensitivity (*Rho* = 0.74, *p* < 0.05), and AD with discrimination (*Rho* = 0.67, *p* < 0.05). For full report of the significant correlations see Table 2.

#### Microstructural - functional measures

For the ASF, contralateral S1 amplitude was negatively linked to MD (*Rho* = –0.55, *p* < 0.05) and contralateral S1 latency was moderately and positively correlated with MD (*Rho* = 0.56, *p* < 0.05) and AD (*Rho* = 0.69, *p* < 0.05). Contralateral S2 amplitude also negatively correlated with MD (*Rho* = –0.60, *p* < 0.05) and AD (*Rho* = –0.55, *p* < 0.05). S2 latency, however, did not show a significant correlation with ASF metrics. Regarding interhemispheric balance, higher diffusivity within the ASF was associated with reduced functional lateralization in S2. Specifically, MD (*Rho* = –0.71, *P <* 0.05), AD (*Rho* = –0.64, *P <* 0.05), and RD (*Rho* = –0.68, *P <* 0.05) each showed strong negative correlations with the S2 lateralization index. No significant correlation was observed between ASF diffusion metrics and S1 lateralization index.

For SCF, contralateral S1 amplitude was strongly correlated with the MD (*Rho* = –0.77, *P <* 0.05), AD (*Rho* = –0.78, *P <* 0.05), and RD (*Rho* = –0.76, *P <* 0.05). Contralateral S2 amplitude showed weaker associations with SCF diffusivity metrics: it was negatively associated with MD (*Rho* = –0.66, *P <* 0.05), AD (*Rho* = –0.58, *P <* 0.05), and RD (*Rho* = –0.67, *P <* 0.05). No significant correlations were observed between S1/S2 latency and SCF diffusivity metrics. Lastly, S1 lateralization was inversely related to AD (*Rho* = –0.72, *P <* 0.05). Similarly, S2 lateralization was negatively correlated with RD (*Rho* = –0.62, *P <* 0.05).

## Discussion

By capitalizing on advanced multimodal neuroimaging, we show that children with HCP exhibit distinct structural and functional neuroplasticity patterns based on their lesion type. Early PV lesions primarily affect the white matter, sparing cortical structures, and thereby allowing for greater potential for adaptive neuroplasticity during development, which results in relatively preserved sensorimotor function. In contrast, late CSC lesions involve direct damage to both cortical and subcortical structures, thereby disrupting essential somatosensory regions and substantially limiting the capacity for adaptive neuroplasticity; severe deficits in sensorimotor outcomes manifest these disruptions. Our findings provide new insights into the neuroplasticity mechanism in HCP and highlight potential targets for optimizing therapeutic interventions.

### CSC and PV lesions cause distinguishable structural damage

CSC lesions disproportionately affected the gray matter and resulted in more extensive volume loss. Most CSC lesions, caused by MCA stroke, affected pre- and post-central gyri, damaging both S1 and S2. One of the key consequences of this structural damage to S1 and S2 was the suppressed SER in the MA hemisphere. This may be due to neuronal necrosis caused by hypoxic-ischemic injury.^44^ In addition, larger lesions, especially in the CSC group, were linked to diminished ability in touch sensitivity and discrimination.

The effects of the insult to specific brain structures were reflected in outcomes as well. Damage to gray matter (i.e., neuronal necrosis) in CSC lesions may eliminate both intra-regional information processing (in S1 and S2) and inter-regional information transfer (across ASF and SCF). These structural damages may disable the processing of afferent information to S1 (also represented as suppressed SER amplitude) and disable the transmitting capabilities of the sensorimotor network (represented as delayed SER).

Somatosensory behavioral results can also be utilized to better contrast between effects of cortical vs. subcortical gray matter damage. PV lesions with minimal (or no) cortical damage showed minimal touch sensitivity deficits; this is possibly due to close to normal low-order sensory processing in S1. Whereas CSC lesions led to severely diminished, or in some cases, complete lack of touch sensitivity. Both lesion types showed deficits in discrimination, more severe in the CSC group. This can be associated with prevalent damage to the thalamus and basal ganglia, especially in the CSC group.^45–47^ Poor (or complete) lack of discriminability in the CSC group can also be associated with the lesion damaging the S2, a crucial hub in the high-order somatosensory network.^48^

### CSC and PV lesions cause distinguishable damage to ASF and SCF

Further illustrating the neuroplasticity potential of children with HCP, tractography showed that ASF can bypass the lesion, projecting to corresponding cortical areas in the MA hemisphere of all children with PV and most children with CSC lesions. Furthermore, microstructural analysis of ASF revealed unilateral damage in the MA hemisphere of both groups, with MD and AD being higher, and more pronounced damage in the CSC group. Increased MD may be caused by gliosis and microscopic cystic degeneration,^49^ while increased AD may indicate lower axonal density and axonal loss.^50^ As a measure of water diffusion perpendicular to fibers, RD was also elevated, particularly in the CSC group, implying possible demyelination, cerebral edema, or myelin pathology.^51^ These diffusivity values in the MA hemisphere of the CSC group were associated with poor touch sensitivity. This relationship highlights the role of ASF, as the first transmitter of afferent inputs, and its axonal integrity in lower-order somatosensory processing. In contrast, ASF diffusivity metrics were not linked to two-point discrimination. This distinction suggests that higher-order somatosensory functions involve more complex and distributed processing pathways, making them less directly tied to afferent tract integrity alone.

Similar inter-group variations in structural integrity were observed in the SCF, with disruptions evident in the MA hemisphere of the CSC group and bilaterally in the PV group. Radiological evaluations also indicated more prevalent damage to the posterior CC body in the PV group. This pattern suggests that the bilateral rise in MD, AD, and RD in the PV group likely indicates arrested SCF maturation, possibly caused by damaged myelination following early prenatal injury.^52^ Higher diffusivity metrics were also linked to both low- and high-order sensory impairments which may implicate the involvement of interhemispheric pathways in somatosensory processing.^46,53^

Previous studies have shown a relationship between CC structural integrity and outcome.^54^ These studies also reported that HCP children with lower levels of CC damage possess a higher potential for upper-extremity function improvement following rehabilitation.^55^ This highlights the importance of preserved interhemispheric connectivity for higher-order somatosensory processing.^56^ In our cohort, disruptions in interhemispheric integration appeared to reflect distinct lesion-related patterns of white matter damage. CSC lesions cause more severe disruption to ASF and SCF fibers in the MA hemisphere. On the other hand, PV lesions primarily affect the ASF unilaterally and SCF bilaterally. The developmental timing of these lesions may explain this pattern. PV lesions precede the ASF development therefore allowing these fibres to bypass the lesion and thus lessen the damage degree. In addition, since CC fibers develop before ASF,^57^ PV lesions are more likely to occur during SCF development which leads to damages to CC and bilateral SCF. Contrarily, CSC lesions occur during (or after) ASF development and after SCF development, increasing the fibers’ susceptibility to direct damage. These findings highlight how lesion type and timing shape somatosensory outcomes in HCP, with late CSC lesions causing direct cortical damage and greater disruption to both afferent processing and intra- and inter-hemispheric connectivity. In contrast, early PV lesions often spare cortical areas and permit adaptive reorganization, particularly for lower-order functions.

### CSC lesions disrupt afferent signal transmission and processing

S1 was localized in the post-central gyrus of the MA hemisphere in most HCP participants. In two participants of the CSC group, the peak amplitude of the first SER component was localized in the subcortical gray matter, near the lesion. These two participants also showed a lack of touch sensitivity and two-point discrimination. Evoked activity in contralateral S1 was suppressed in both hands of the CSC group and the NPH of the PV group. These observations contrast with the expected diminished S1 amplitude in response to only the PH stimulation. Instead, they possibly reveal a disruption in the mediation of information flow and processing in S1.^58^ This pattern may reflect that damage to deep structures leads to suppressed S1 activity in both hemispheres of children with CSC lesion.^59^ In contrast, S1 amplitude in the MA hemisphere of the PV group was comparable to TD. Although unexpected, this finding may reflect functional neuroplasticity and the compensatory engagement of spared cortex to offset the disrupted input and damage to the ASF.^58^

A robust association was found between S1 amplitude responses and ASF/SCF diffusion metrics. In ASF, MD was correlated negatively with contralateral S1 amplitude, possibly implying that axonal loss (or demyelination) compromises the afferent fibers’ capacity to transmit action potentials and activate S1, thereby reducing cortical responsiveness. In SCF, MD, AD, and RD were correlated negatively with S1 amplitude. These associations reveal a link between S1 activity— as the first cortical hub for lower- and higher-order somatosensory processing— and commissural fibers’ structural integrity.^60^ Reduced S1 amplitude was associated with elevated touch sensitivity and two-point discrimination threshold; this indicates that diminished neuronal activation in S1 is tied to impaired sensory processing.^61^ This was particularly evident for the PH of children with CSC lesions, where evoked responses were often severely attenuated and behavioral performance was poorest.

The prolonged S1 latency in the MA hemisphere of the CSC group likely reflects impaired conduction through ASF due to disrupted microstructural integrity. These delayed S1 responses were strongly associated with elevated MD and AD in these fibers. This reinforces the interpretation that compromised axonal density and myelination slow action potential transmission and degrade the temporal dynamics of cortical activation.^62^ Notably, the latency of contralateral S1 responses also correlated with higher sensory thresholds. These delays likely reduce the precision of sensory processing timing, contributing to diminished perceptual acuity.^61^ However, when examining higher-order sensory processing, the complexity and heterogeneity of parallel information pathways introduce considerable variability.^58^ As a result, the latency of second peaks in cortical evoked responses showed high variance across all subjects.

### CSC lesions disrupt the interhemispheric balance

We have previously reported ipsilateral S1 and S2 activities in response to haptic stimulation in TD children.^63^ These activities were less prominent than those in the contralateral hemisphere, establishing an “interhemispheric balance” in typical somatosensory processing. Here, we quantified this balance with SLIs of S1 and S2. TD children exhibited high SLI in S1 bilaterally, reflecting strong lateralization and minimal involvement of the ipsilateral hemisphere in low-order somatosensory processing. In contrast, the CSC group showed lower SLI in S1 in response to PH stimulation. This suggests increased involvement of ipsilateral S1 and a shift in early sensory processing. In S2, TD children demonstrated lower SLI consistent with typical bilateral involvement in high-order processing.^47^ The abnormally low SLI in S2 during PH stimulation in the CSC group suggests disrupted interhemispheric equilibrium and heightened contribution of contralesional S2, possibly due to reorganization of sensory processing pathways.^64^

Diffusivity metrics were inversely associated with SLIs in both S1 and S2. Higher MD, AD, and RD in ASF were linked to S2 SLI. This is due to impaired afferent signal transmission, possibly disrupting serial and parallel somatosensory processing in S2 bilaterally.^63^ In addition, alterations in SCF diffusivity parameters were related to decreased S1 and S2 lateralization. This suggests that white matter injury, particularly in the commissural fibers, may drive the atypical bilateral processing observed in our functional analyses, further linking microstructural damage to maladaptive reorganization. Another possible cause for the effect of SCF damage on imbalanced SLI is the disrupted inhibitory role of the ipsilateral S1 in somatosensory processing.^65,66^

Reduced SLI in S1 and S2 correlated strongly with behavioral deficits. Lower SLIs— indicating greater reliance on ipsilateral or bilateral processing—were associated with impaired discriminability.^64^ This pattern was particularly evident in the CSC group, suggesting that a shift away from typical contralateral dominance (interhemispheric balance) may reflect maladaptive reorganization that fails to support functional recovery. The strengthened involvement of contralesional S2 after CSC lesions might reflect the emergence of potential alternative pathways in the absence of ipsilesional S2. Yet, a prevalent interhemispheric dissociation between S1 and S2 in response to afferent somatosensory inputs limits this potential. These findings demonstrate that CSC lesions disrupt typical interhemispheric balance by reducing S1 and S2 lateralization and increasing bilateral activation, linking microstructural white matter damage to maladaptive sensory reorganization and poorer discrimination. Ultimately, the observed bilateral somatosensory representation in the reorganized brain of children with CSC lesions may impact treatment decisions, especially regarding direct invasive or non-invasive brain stimulation targets.

### Limitations and Future Directions

Due to the heterogeneity and lower prevalence of other etiologies in HCP (different brain malformations), we were not able to classify these participants in a coherent group. Furthermore, the small sample size, especially in the CSC group, may limit the generalizability of our findings. A comprehensive study with a larger sample size may address these limitations. Due to the cross-sectional design, we also cannot directly assess developmental trajectories of neuroplasticity. Longitudinal studies are needed to explore how these functional and structural patterns evolve with age and intervention. Finally, further exploration of secondary somatosensory areas (e.g., S2 connectivity) and cortical-subcortical interactions may provide additional insights into compensatory mechanisms.

## Conclusion

This study elucidates distinct neuroplasticity profiles in children with CSC and PV lesions. CSC lesions are characterized by more severe cortical and subcortical damage, with limited compensatory reorganization. In contrast, PV lesions involve less direct cortical injury, allowing for more effective structural and functional adaptation. Notably, pronounced contralesional activation of S1 and S2 in response to haptic stimulation of the PH of the CSC group suggests some retained neuroplasticity potential. These findings underscore the importance of early detection and intervention—such as somatosensory training—for children with CSC lesions to improve sensory and motor outcomes. Altogether, our findings highlight the importance of lesion-specific treatment plans and the benefit of combining structural and functional neuroimaging to guide tailored rehabilitation programs. These revelations deepen our knowledge of adaptive neuroplasticity in pediatric HCP and open the path for enhancing sensorimotor results utilizing precision medicine strategies.

## Data availability

The data are available from the corresponding author upon request.

## Data Availability

All data produced in the present study are available upon reasonable request to the authors.

## Acknowledgement

We thank Dr. Sabrina Shandley and Eryn Armstrong for their contribution in behavioral assessment.

## Authors Contributions

S.S., Y.S., and C.P. contributed to the conception and design of the study. S.S., Y.S., A.V., S.S., H.R., and M.A.G.M contributed to the acquisition and analysis of the data. S.S., M.B., F.A., B.K., and W.M. contributed to diagnosis, lesion type determination, and clinical assessments. S.S., W.M., and C.P. contributed to drafting a significant portion of the manuscript and figures.

## Supplementary material

Supplementary material is available online.

