## Supplemental Table 1 for "Functional and Structural Neuroplasticity of Somatosensory System in Hemiplegic Cerebral Palsy"

**Table S1 Behavioral, functional, and structural results for the 'other' lesion type**

| | | #10 | | #11 | | #16 | |
|---|---|---|---|---|---|---|---|
| **Metric** | | **NPH** | **PH** | **NPH** | **PH** | **NPH** | **PH** |
| **Behavioral** | **Touch sensitivity (g)** | 0.07 | 0.07 | 0.07 | 0.16 | 0.07 | 0.16 |
| | **Two-point discrimination (mm)** | 2 | 2 | 2 | 3 | 3 | 10 |
| | **Range of motion (%)** | 100 | 92.6 | 100 | 90.6 | 100 | 83.3 |
| | **Accuracy (%)** | 100 | 100 | 100 | 96.3 | 100 | 86.4 |
| | **Dexterity (%)** | 100 | 75 | 100 | 93.8 | 100 | 38.5 |
| | **Fluency (%)** | 100 | 85.7 | 100 | 76.2 | 100 | 66.7 |
| **Functional** | **Contra S1 amp (AU)** | 0.72 | 0.17 | 0.98 | 0.39 | 0.51 | 0.85 |
| | **Contra S1 latency (ms)** | 65 | 75 | 58 | 57 | 89 | 86 |
| | **S1 LI** | 0.7 | 0.98 | 0.68 | -0.11 | 0.70 | 0.23 |
| | **Contra S2 amp (AU)** | 1.02 | 0.39 | 0.38 | 0.97 | 0.68 | 0.37 |
| | **Contra S2 latency (ms)** | 174 | 142 | 111 | 136 | 140 | 141 |
| | **S2 LI** | 0.49 | 0.09 | -0.2 | 0.89 | 0.57 | 0.11 |
| | | **LA** | **MA** | **LA** | **MA** | **LA** | **MA** |
| **Structural** | **ASF MD** | 0.79 | 0.82 | 0.76 | 0.77 | 0.86 | 1.03 |
| | **ASF AD** | 1.29 | 1.35 | 1.18 | 1.30 | 1.33 | 1.49 |
| | **ASF RD** | 0.53 | 0.55 | 0.55 | 0.51 | 0.62 | 0.79 |
| | **SCF MD** | 0.81 | 0.75 | 0.89 | 0.75 | 1.28 | 1.20 |
| | **SCF AD** | 1.34 | 1.29 | 1.35 | 1.23 | 1.68 | 1.67 |
| | **SCF RD** | 0.55 | 0.48 | 0.66 | 0.51 | 1.09 | 0.96 |

AD = axial diffusivity; amp = amplitude; ASF = ascending sensory fiber; LA = less affected; LI = lateralization index; MA = more affected; MD = mean diffusivity; NPH = non-paretic hand; PH = paretic hand; RD = radial diffusivity; S1 = primary somatosensory cortex; S2 = secondary somatosensory cortex; SCF = somatosensory commissural fiber.
